# A genome-wide association study of hidradenitis suppurativa from the VA’s Million Veteran Program

**DOI:** 10.1101/2025.06.23.25330121

**Authors:** Zachary Wendland, Craig C. Teerlink, Kathryn M. Pridgen, Sydney Lo, Christopher Sayed, KR Van Straalen, Catherine Tcheandjieu, Philip S. Tsao, Kyong-Mi Chang, Yun Li, Karen L Mohlke, Quan Sun, Julie A. Lynch, Noah Goldfarb

## Abstract

**Background:** Data from family and twin studies as well as prior genome-wide association meta-analyses suggest that hidradenitis suppurativa (HS) has a hereditary component.

**Objective:** Identify genetic variants associated with HS.

**Methods:** A case-control genome-wide association study (GWAS) was performed on participants with a diagnosis of HS defined as at least one instance of ICD9 705.83 or ICD10 L73.2 from participants in the VA’s Million Veteran Program.

**Results:** 4,959 participants with HS were identified among 597,819 MVP participants. The multi-population GWAS identified two significant loci associated with HS, including a novel HS-related variant on chromosome 6 near *HLA-DRB1* (lead variant rs679242), and confirmed a previously identified locus on chromosome 17 near *SOX9* (rs55811634). The following previously identified loci achieved suggestive evidence for association (p<1x10^-3^): rs17090189 (near *KLF5*), rs121908120 (2q35), rs10816701 (9q31.3), rs17090189 (13q22.1), and rs17103088 (14q24.3).

**Conclusion:** The analysis of the MVP resource for HS identified a novel signal on chromosome 6 near *HLA-DRB1*, and identified significant evidence and suggestive evidence for several previously reported signals for HS.

## Introduction

Hidradenitis suppurativa (HS) is a chronic, debilitating inflammatory skin disorder characterized by painful nodules, abscesses, and tunnels predominantly affecting intertriginous areas of the body.^1^ HS is associated with comorbidities across multiple body systems, including rheumatologic, metabolic and cardiovascular diseases (CVD).^2^ The association is strong enough that guidelines recommend screening for hypertension, diabetes mellitus, metabolic syndrome and inflammatory arthritis in patients with HS.^3^ The recent creation of a polygenic risk score for HS using data from FinnGen showed that HS is genetically associated with an increased risk for diabetes mellitus and coronary artery disease.^4^ This finding has also been corroborated through mendelian randomization studies suggesting possible causal relationships between HS and cardiovascular disease.^5–7^

HS has a genetic predisposition, with twin studies demonstrating 77-80% heritability.^8,9^ In some families, this genetic predisposition is fairly strong with autosomal dominant inheritance patterns.^8^ Overall, 30-40% of patients with HS report at least one first degree relative with HS.^9,10^ While 5-6% of patients with HS have been found to have causative loss-of-function mutations in the gamma-secretase complex, the majority of patients with HS do not have an identified genetic variant.^8,11^ Through two large genome-wide association studies, several genetic variants associated with HS have been identified.

Sun et al.^12^ performed a multi-ancestry GWAS including 720 participants with HS from the University of North Carolina (ProCare Registry) in combination with data from UK Biobank, FinnGen, and BioVU, which identified single-nucleotide polymorphism (SNP) variants on chromosome 13 near *KLF5* and on chromosome 17 near *SOX9*.^12^ Anderson RK et al combined five data sets from Denmark, Finland, Iceland, as well as the United Kingdom and United States, including 4,814 participants with HS.^13^ This meta-analysis confirmed a previous finding (the variant near *KLF5*) and identified 7 novel variants on chromosome 1 near *NCSTN*, chromosome 2 near *WNT10A,* chromosome 14 near *TMED10,* chromosome 19 near *PSENEN* and three other variants on chromosomes 6, 9, and 17.

Building upon these promising findings, we conducted a GWAS in a large, diverse cohort of patients with HS enrolled in the VA’s Million Veteran Program (MVP) with the intent of validating prior variants, and if possible, identifying additional associations. Our study aimed to provide a comprehensive and detailed exploration of the genomic landscape of HS within this unique population, with the ultimate goal of advancing our understanding of the genetic factors contributing to HS.

## Methods

### Million Veteran Program

The MVP, a genetic repository of US Veterans linked to clinical records from a nationwide electronic health record system, was launched in 2011 by the Veterans Health Administration (VHA) Office of Research and Development and received ethical and study protocol approval from the VA Central Institutional Review Board.^14^ At the time of our study, approximately 650,000 MVP participants had available genotype data on a customized Affymetrix Axiom biobank array (MVP 1.0 Genotyping Array) which contains 723,305 variants, enriched for low-frequency variants in those of African and Hispanic ancestry.^15^ Details on imputation and quality control have been described previously.^16^ Our analysis used Release 4 of MVP imputed from approximately 97,000 background genomes in TOPMed.^17^ We also use 102,677 whole genomes from MVP Release2 of whole-genome sequencing (https://va-big-data-genomics.github.io/jekyll/update/2023/11/17/Data-Release-2-process-summary.html) to confirm imputed genotype calls for variants in the HLA region.

### Study subjects

MVP participants were identified as having HS with at least one instance of an HS diagnostic code of ICD9 705.83 or ICD10 L73.2, which is validate case-finding algorithm with 100% sensitivity and 83% specificity.^18^ Demographic and clinical information were collected from Corporate Data Warehouse and comorbidities were identified using diagnostic codes and presented with summary statistics. Differences between HS and non-HS cohorts were analyzed with two-sided t-tests for continuous variables and Chi-squared tests for categorical variables (R v4.2.1). P<0.05 was considered statistically significant.

### Genome-wide association analysis

In each ancestry, we tested roughly 15.5 million (M) imputed SNPs that passed quality control (i.e., HWE >1x10^-10^, INFO >0.3) with minor allele frequency (MAF) greater than 0.01 for association with HS through logistic regression assuming an additive model of variants using REGENIE software using Firth regression for variants with p<0.01.^19^ Covariates included age at enrollment in MVP, sex, and the first ten principal components of genetically-inferred ancestry. We also performed a multi-ancestry meta-analysis in a fixed-effects model using METAL software^20^ with inverse-weighting of the effect estimates. SNPs were considered genome-wide significant if they passed the conventional threshold of p<5x10^-8^.

### Secondary signal analysis

Independent signals at regions with multiple significant SNPs were evaluated by individual-level conditional association regression analysis by introducing the lead SNP in the genetic region (defined as a 50-Kb flanking region of significant markers) as an additional covariate and re- analyzing the region. SNPs with p<5x10^-8^ were considered independent signals.

### Putative causal variant mapping

All imputed variants in MVP were annotated with ANNOVAR software^21^ and RegulomeDB.^22^ We extracted all predicted loss-of-function and missense variants as well RegulomeDB score for non-coding variants. Linkage disequilibrium (LD) with established variants was calculated via PLINK 2.0^23^ for multi-ancestry, European ancestry (EA), African ancestry (AA), and Hispanic ancestry (HA) lead SNPs based on internal estimates using the MVP dataset for the respective underlying population. For the multi-ancestry coding variants, we used the EA panel for LD calculation. Coding variants or high-scoring non-coding variants that were in strong LD (r^2^>0.7) with lead SNPs and had a moderately strong statistical association (p<1x10^-5^) were considered the putative causal drivers of the observed associations. Pairwise LD between lead SNPs and other significant SNPs was estimated using LDlink.^24^

### Phenome-wide association study of lead SNPs

Lead SNPs from the meta-analysis were investigated in several external resources that provide phenome-wide associations (PheWAS), including UKBiobank (via PheWeb),^17^ HugeAMP (https://hugeamp.org), and FinnGen (data freeze 9).^25^

### Novel variant confirmation

Novel lead SNP(s) in the VA study were previously analyzed in a meta-analysis including HS Program for Research and Care Excellence (HS ProCARE) from the University of North Carolina Department of Dermatology,^12^ UKBiobank, and FinnGen; we consulted results from this analysis to confirm findings from this study.

## Results

### Study subjects

We identified 4,959 (0.83%) participants (3,967 [80%] males; 992 [20%] females) with at least one validated^18^ instance of an HS diagnostic code (ICD9 code 705.83 or ICD10 code L73.2) (**Table 1**). Of these subjects, 1,931 were AA, 2,613 were EA, and 415 were HA, among 597,819 MVP participants of the same ancestral origins (AA: 112,279; EA: 437,982; HA: 47,558) (**Table 1**). This case search strategy has been previously validated by Strunk et al. (2017).^18^ The HS cohort was younger than controls, with increased prevalence of obesity and tobacco use (**Table 1**).

**Table 1.** Demographics of hidradenitis suppurativa (HS) and non-HS cohorts in the VA Health Care System

|  | HS<br>n = 4,959 | Non-HS<br>n = 592,860 | p-value |
| --- | --- | --- | --- |
| Age, mean (SD) | 55.3 (12.6) | 62.0 (14.0) | p < 0.001 |
| Male sex, n (%) | 3967 (80.0%) | 474,177 (91.3%) | p < 0.001 |
| Race/ethnicity, n (%) |  |  | p < 0.001 |
| African American (%) | 1,931 (38.9%) | 110,348 (18.6%) |  |
| European American (%) | 2,613 (52.7%) | 435,369 (73.4%) |  |
| Hispanic American (%) | 415 (8.4%) | 47,173 (8.0%) |  |
| BMI, mean (SD) | 33.9 (7.2) | 31.3 (6.2) | p < 0.001 |
| Never tobacco use, n (%) | 550 (11.1%) | 112,051 (18.9%) | p < 0.001 |

### Genome-wide association

The population-specific GWAS identified no significant associations in the AA population, one locus in the EA population (rs10889867 at 1p31.1 [*ANKRD13C*]), and two loci in the HA population ([no rsID] at 6q25.3 [*FNDC1*], and rs72755927 at 9q31.3 [*ACTL7B*]), although these population-specific findings were not included in the meta-analysis. (Supplemental Figures 1-3) The meta-analysis did identify two other regions: rs679242 at 6p21.32 (*HLA-DRB1*) and rs55811634 at 17q24.3 (*SOX9*). (Figure 1) **Table 2** shows lead SNPs for significant markers in the respective populations and the meta-analysis. No secondary signals were identified in any of the populations or the meta-analysis.

**Figure 1.**
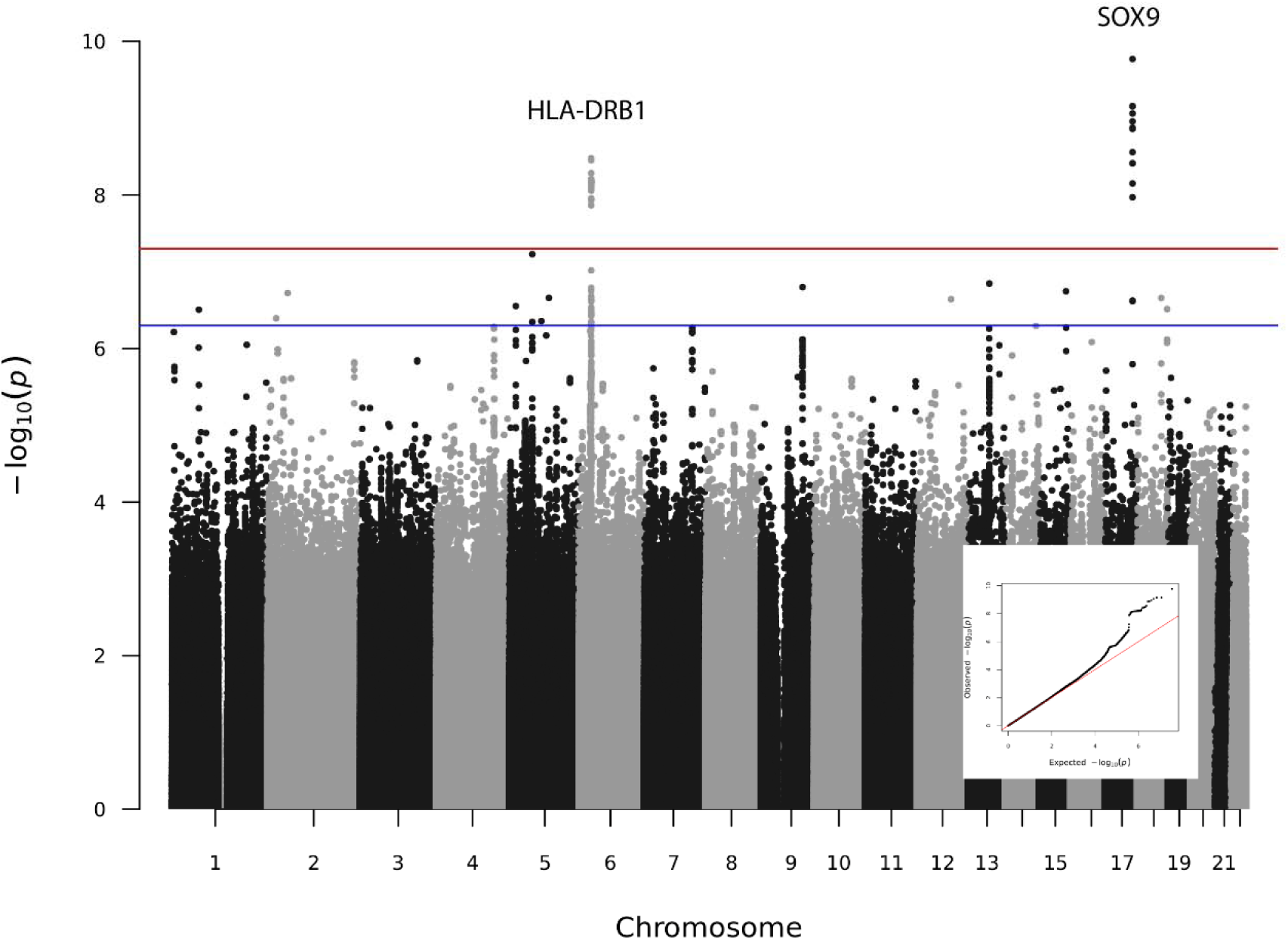
GWAS meta-analysis of genetic variants from the Million Veteran Program in participants with hidradenitis suppurativa

**Table 2.**
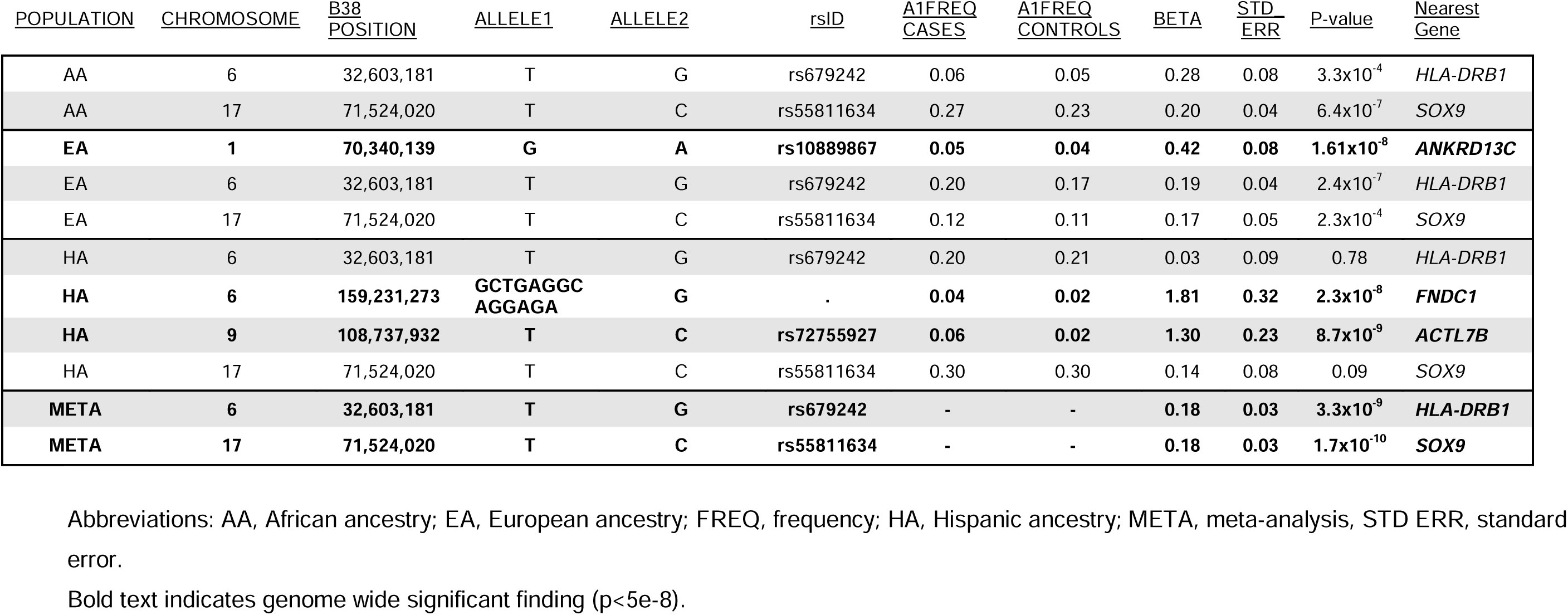
Lead SNPs from genome-wide association by population and meta-analysis

### Putative causal variant mapping

In the EA population, the lead SNPs at 1p31.3 (*CACHD1*; rs6588100) and 1p31.1 (*ANKRD13C*; rs10889867) were the only significant SNPs in the region and are considered the putative causal variants for those regions. In the HA population, the lead SNP at 6q25.3 (*FNDC1*; no rsID) was the only significant SNP and is considered the putative causal variant. Both significant SNPs at 9q31.3 (*ATCL7B*; rs72753987 and rs72755927) are non-coding with high RegulomeDB scores (indicating low potential for regulation) and are equally plausible to contribute to HS. In the meta-analysis, all SNPs in the 6p21.32 (near HLA-DRB1) are non-coding, and 19 of the 35 significant SNPs have a RegulomeDB score of 1b or 1f (indicating high potential for regulation), including the lead SNP rs679242. Similarly, in the 17q24.3 (*SOX9*) region, all significant SNPs are non-coding with mid-range (4-7) RegulomeDB scores (indicating low to moderate potential for regulation). Both regions show multiple variants in tight linkage disequilibrium.

### Phenome-wide association study of lead SNPs

We did not identify any significant PheWAS results for the population-specific variants. Meta-analysis SNP rs679242 (6p21.32; *HLA-DRB1*) has been previously significantly associated with several diseases including rheumatoid arthritis (p=1.1x10^-121^), type 1 diabetes (p=4.6x10^-63^), and type 2 diabetes (p=2.8x10^-14^) (**Table 3**). Diabetes mellitus types I and II were significantly increased in our HS cohort compared with non-HS participants across ancestries (p<0.001).

**Table 3.** Results of PheWAS studies from three large external databases

| Chr | rsID | Nearest Gene | UKBiobank | AMP (Metabolic Disease Portal) | FinnGen |
| --- | --- | --- | --- | --- | --- |
| 6 | rs679242 | qter *HLA-DRB1* | Rheumatoid arthritis ($p=1.1 \times 10^{-121}$)<br>Type 1 diabetes ($p=4.6 \times 10^{-63}$)<br>Celiac disease ($p=6.6 \times 10^{-54}$)<br>Asthma ($p=2.7 \times 10^{-45}$)<br>Hypothyroidism ($p=1.1 \times 10^{-35}$)<br>Nasal polyps ($p=1.7 \times 10^{-28}$)<br>Polymyalgia rheumatica ($p=2.7 \times 10^{-25}$)<br>Diabetic retinopathy ($p=6.2 \times 10^{-20}$)<br>Type 2 diabetes ($p=2.8 \times 10^{-14}$)<br>Multiple sclerosis ($p=6.9 \times 10^{-13}$) | Rheumatoid arthritis ($p=5.0 \times 10^{-324}$)<br>Type 1 diabetes ($p=5.0 \times 10^{-324}$)<br>Neutrophil count ($p=4.1 \times 10^{-118}$)<br>White blood cell count ($p=2.4 \times 10^{-102}$)<br>Eosinophil count ($p=6.8 \times 10^{-23}$)<br>Type 2 diabetes ($p=4.0 \times 10^{-18}$)<br>HDL cholesterol ($p=2.2 \times 10^{-16}$)<br>SLE ($p=1.7 \times 10^{-15}$)<br>Alkaline phosphatase ($p=1.4 \times 10^{-13}$)<br>eGFR-creat ($p=2.0 \times 10^{-13}$) | Type 1 diabetes ($p=5 \times 10^{-324}$)<br>Rheumatoid arthritis ($p=5 \times 10^{-324}$)<br>Diabetic retinopathy ($p=5 \times 10^{-324}$)<br>Diabetic maculopathy ($p=3.1 \times 10^{-170}$)<br>Hypothyroidism ($p=3.3 \times 10^{-80}$)<br>Type 2 diabetes ($p=4.3 \times 10^{-65}$)<br>Vitreous bleeding ($p=1.1 \times 10^{-62}$)<br>Glomerular disorders ($p=5.8 \times 10^{-49}$)<br>Polymyalgia rheumatica ($p=5.5 \times 10^{-42}$)<br>Asthma ($p=4.1 \times 10^{-29}$) |
| 17 | rs55811634 | pter *SOX9* | (no significant results) | (no significant results) | Hidradenitis suppurativa ($p=2.4 \times 10^{-8}$) |

Top 10 results reported with $p < 5 \times 10^{-5}$ as of March 10, 2023
Abbreviations: Chr, chromosome; SLE: systemic lupus erythematosus; eGFR-creat: estimated glomerular filtration rate-creatinine; high density lipoprotein

This corresponded to a statistically significant increased risk of cardiometabolic conditions in AA and EA participants within our dataset (p<0.05) (Supplemental Table 1). The SNP rs55811634 (17q24.3; *SOX9*) has been previously associated with HS.^12^

### Confirmation of SNPs

Two previously reported SNPs^12^ have been significantly associated with HS: rs10512572 (near *SOX9*), which met genome-wide significant in MVP meta-analysis, and rs17090189 (near *KLF5*), which had p=0.0001 (suggestive evidence) in the MVP meta-analysis; therefore, our analysis of the MVP cohort statistically confirmed one of these SNP’s association with HS. A previous meta-analysis conducted for HS^13^ identified several novel associations. Analysis of these SNPs in MVP identified significant evidence (p<5x10^-8^) for the previously reported SNP at *SOX9* (chr17:71519473:A:G [p=8.7e^-10^]) and suggestive evidence (p<1x10^-3^) for SNPs rs121908120 (chr2:218890289:T:A; p=6.0e^-4^), rs10816701 (chr9:108703801:C:T; p=1.7e^-6^), rs17090189 (chr13:73432270:A:G; p=1.2e^-4^), and rs17103088 (chr14:75230213:A:G; p=1.0e^-3^).

The lead variant (rs679242) identified on chromosome 6 near *HLA-DRB1* in this study was confirmed at the marginal p<0.05 level in a meta-analysis of two large biobanks, FinnGen and UK Biobank, as well as among the participants in HS ProCARE. (Supplemental Table 2).

Given the potential for inaccuracy of variant calls in the HLA region when relying on imputed genomes,^26^ we checked the concordance of rare allele carriage between the imputed genomes and whole genome sequences for ∼102K subjects who had both data types available for rs679242. The concordance rate of the rare allele was 99.93% between these data types indicating that the genotypes in the imputed dataset were accurately called.

## Discussion

The comprehensive GWAS conducted within the VA’s MVP provides valuable insights into our understanding of genetic drivers of HS pathogenesis and associated comorbidities. The meta-analysis herein identified HS-associated loci at 6p21.32 (*HLA-DRB1*; lead SNP rs679242) and 17q24.3 (*SOX9*; lead SNP rs55811634) and confirmed the lead SNP rs17090189 (near *KLF5*). Given the tight linkage disequilibrium in both the 6p21.32 (near *HLA-DRB1*) region and the 17q24.3 (*SOX9*) region, each set of SNPs are likely inherited together and one or more variants may contribute to risk in their respective region.

*HLA-DRB1* is the gene encoding one of the class II human leukocyte antigen (HLA) beta chains. HLA molecules are transmembrane glycoproteins found on nearly all nucleated cells in the body, playing a crucial role in immune regulation; specifically, class I and II MHC molecules are responsible for presenting peptides to CD8 cytotoxic cells and CD4 cells, respectively.^27^ Class II genes (*HLA-DR, DQ,* and *DP*) produce the alpha (A1) and beta chains (B1-4) of the corresponding HLA class II molecule found on antigen-presenting cells responsible for presenting extracellular pathogens to T-cells resulting in an immune response.^28^

*HLA-DRB1* is associated with several autoimmune and autoinflammatory disorders, most notably rheumatoid arthritis (RA). Patients with HS have been found to have a three-fold increased risk of inflammatory arthritis, specifically spondylarthritis.^29^ In the recent 2023 comorbidity screening guidelines for HS, inflammatory arthritis had a “B” strength of recommendation supporting physicians screen for inflammatory arthritis in patients with HS.^3^ In the meta-analysis, SNP rs679242 (*HLA-DRB1*) was significantly associated (p<5e-8) with RA in three large databases (UKBiobank, HugeAMP, and FinnGen), and reported in several prior studies.^28,30–32^ While the precise mechanism linking *HLA-DRB1* variants and RA remains unknown, the “shared epitope hypothesis” suggests that certain alleles with a conserved sequence of five amino acids directly contribute to pathogenesis by permitting the incorrect presentation of autoantigens to T-cells by antigen-presenting cells resulting in autoinflammation or autoimmunity.^28,33,34^ These class II HLA shared epitopes have been found to be associated with protein N-linked glycosylation^35^ and increased tissue citrullination, which has also been identified in HS skin compared to controls.^36^ In HS, colocalization of neutrophil extracellular traps have also been identified at the sites of these citrullinated antigens as well as antibody formation against them.^36^ In addition, *HLA-DRB1* alleles have been found to result in polarization toward M1 pro-inflammatory macrophages, as are seen in HS,^37^ compared to M2 macrophages.

Likewise, multiple studies have demonstrated an association between *HLA-DRB1* and type 1 and 2 diabetes mellitus.^38–41^ This is in accordance with prior studies demonstrating increased risk of diabetes mellitus associated with HS, supports current guidelines recommending screening for diabetes mellitus in HS populations.^3^ The association of *HLA-DRB1* with diabetes mellitus 1 and 2 in all three large databases (UKBiobank, HugeAMP, and FinnGen), in addition to the association with aberrant high density lipoprotein (HDL) levels in HugeAMP, may partly explain the increased risk cardiovascular events and mortality associated with HS.^3^ Interestingly, genetic variants of *HLA-DRB1* have been identified as key genes that predispose RA patients to CVD and adverse events.^42–45^

In addition to its association with several HS comorbidities*, HLA-DRB1* has also been identified as one of 27 genes that is hypermethylated in peripheral blood of patients with HS.^46^ This could indicate that these gene variants near HLA-DRB1 in non-coding regions of DNA may be regulators of epigenetic methylation. In a study utilizing the UK Biobank to evaluate the relationship between HLA alleles and HS risk across various Fitzpatrick skin types, eighteen HLA variants conferred increased risk or protective affects for HS, but only HLA-DRB1*01:01 demonstrated a statistically significant protective effect in all skin types.^47^ This is supported by prior smaller studies showing significant differences in various HLA allele frequencies amongst patients with HS.^48,49^ In addition, both methylation studies from the blood of patients with HS.^50^ and proteome profiles from the blood of patients with polygenic risk for HS^4^ demonstrate HLA presentation pathways and proteins, respectively, are some of the most enriched or altered compared to controls. While several HLA proteins seem to influence genetic risk for HS that differ across skin types, we postulate that genes that regulate HLA-DRB1 transcription are likely the most common among patients with HS across ethnic and racial groups to meet genome-wide significance in this meta-analysis.

Consistent with the findings in prior literature,^12^ we identified the variant near *SOX9* as significant within the MVP cohort of patients with HS. The variant near *SOX9* has not yet been linked to any quantitative trait locus effects.^12,13^ However, *SOX9* encodes SRY-box transcription factor 9 (SOX9) stem cell transcription regulator and is expressed in the epidermal basal layer, sebaceous glands, hair follicle stem cells, and outer root sheath (ORS).^51^ SOX9 is important for maintaining hair follicle bulge stem cells that form the ORS through the hair cycle, as well as capability of interfollicular epidermal wound regeneration.^52,53^ Mouse models missing SOX9 in the hair follicles are unable to revert to the quiescent state, eventually resulting in alopecia, as well compromised epidermal wound healing capacity.^52,53^

Conversely, inhibition of SOX9 has been demonstrated to upregulate matrix metalloproteinase (MMP)1, MMP2, and interleukin (IL)-8, which are linked to tumorigenesis and inflammation.^54^ In mouse models, mice lacking SOX9 demonstrate follicular degeneration, epidermal thickening, and subsequent dermal scarring.^54^ Furthermore, upon inhibition of SOX9, a cascade of effects were observed in mice, including formation of keratin pearls, loss of keratin 1 (K1) expression in the upper ORS, ultimately leading to the loss of hair follicle stem cells and regenerative potential.^52^ Histologically this translates to follicular degeneration, epidermal thickening, and subsequent dermal scarring. Interestingly, ORS cells from HS patients also have been found to have increased number of proliferating progenitor cells and loss of quiescent stem cells.^55^ In addition, Notch signaling is required for *SOX9* expression in various cell types.^56–58^ This may be a link from the γ-secretase complex (a protease complex essential for Notch signaling) mutations identified in some HS familial cases to HS pathology.^11^ The role of SOX9 in inflammation pathways is less clear and is likely different in different cell types. For example, in human dental pulp cells, *SOX9* deletion upregulates MMP2 and MMP13 and augments IL-8 in the presence of TNF-α. On the other hand, in cardiac fibroblasts, *SOX9* deletion reduces MMP2, and in hepatic ischemia/reperfusion models, SOX9 augments proinflammatory cytokines TNF-α, IL-1β, IL-6 and TGF-β 1.^59,60^

Although it did not meet genome-wide significant in the MVP study population, rs17090189 (p=2.1x10^-8^) near *KLF5* was confirmed in this cohort as associated with HS, validating previous findings.^12^ Like the variant near SOX9, this variant has not been linked to quantitative trait locus effects.^12,13^ The *KLF5* gene encodes Kruppel-like factor 5 (KLF5), also known as BTEB2, which is a zinc finger transcription factor that is important for regeneration and proliferation for many organ systems.^61^ For skin, it is a regular of epidermal stems cells, found most highly expressed in the basal and suprabasal epidermal layers.^62^ Mice lacking epidermal KLF5 die soon after birth due to defects in barrier function.^63^ In addition, KLF5 is crucial for epidermal lipid biosynthesis and metabolism, especially for sphingolipid. *Klf5*-deficient mice had defects in their lipid envelope with loss of complex sphingolipids and reduced lipid secretory function.^61^ ^63^ HS lesions, similarly, have aberrant sphingolipid metabolism compared to controls. HS lesions have reduced expression of enzymes that generate sphingomyelin and ceramides and increased expression of enzymes that breakdown ceramides.^64^

GWAS studies are in themselves limited to identifying associated variants and do not identify the genes that are responsible for HS risk. While researchers can theorize about the roles of these variants in HS biology, these studies cannot confirm a causal effect or explain the underlying pathogenesis. Functional studies are therefore needed to assess whether the identified variants or the surrounding genes affect HS development. Another limitation of this study is our unique Veteran population, which limits generalizability of this study. In addition, while the diversity of MVP surpasses that of other large biobanks,^16^ it is still composed predominately of persons of European ancestry. Additionally, the VA and MVP population is mostly male, and HS is more common in female individuals, with a reported female-to-male ratio of approximately 3:1.^65^ Although the MVP is extensive, further efforts must be made to include genotype data from underrepresented populations.

In summary, our analysis provided statistical confirmation of two previously reported SNPs significantly associated with HS, rs55811634 (near *SOX9*) and rs17090189 (near *KLF5*).

Additionally, our study identified additional SNPs near *HLA-DRB1,* which is a gene that has been associated with a number of known HS comorbidities. This GWAS underscores the intricate genetic landscape of HS, pointing towards significant genetic markers linked to disease development and coexisting conditions. Future studies are planned to evaluate the functional effects of these HS-associated genes on HS pathology.

## Data Availability

Patient-level data are currently accessible to all VA researchers with appropriate IRB approvals.

## Data Availability

Patient-level data are currently accessible to all VA researchers with appropriate IRB approvals.

## Acknowledgements

This work was supported using resources and facilities of the Department of Veterans Affairs (VA) I01-BX003362 (Chang, Tsao) VA Informatics and Computing Infrastructure (VINCI) VA HSR RES 130457 and NIH R21 AR075996 (Sayed, Li, Mohlke). This publication does not represent the views of the Department of Veterans Affairs or the United States Government.

## Data sharing statement

Patient-level data are currently accessible to all VA researchers with appropriate IRB approvals.

## Ethics statement

The Million Veteran Program received ethical and study protocol approval from the VA Central Institutional Review Board. Informed consent was received from all participants.

## References

1. Sabat, R., Jemec, G.B.E., Matusiak, Ł., Kimball, A.B., Prens, E., and Wolk, K. (2020). Hidradenitis suppurativa. Nat Rev Dis Primers 6, 18. 10.1038/s41572-020-0149-1.

2. Cartron, A., and Driscoll, M.S. (2019). Comorbidities of hidradenitis suppurativa: A review of the literature. Int J Womens Dermatol 5, 330–334. 10.1016/j.ijwd.2019.06.026.

3. Garg, A., Malviya, N., Strunk, A., Wright, S., Alavi, A., Alhusayen, R., Alikhan, A., Daveluy, S.D., Delorme, I., Goldfarb, N., et al. (2022). Comorbidity screening in hidradenitis suppurativa: Evidence-based recommendations from the US and Canadian Hidradenitis Suppurativa Foundations. J Am Acad Dermatol 86, 1092–1101. 10.1016/j.jaad.2021.01.059.

4. Nielsen, V.W., Bundgaard Vad, O., Holgersen, N., Paludan-Müller, C., Meseguer Monfort, L., Beyer, A.F., Jemec, G.B.E., Kjærsgaard Andersen, R., Egeberg, A., Thyssen, J.P., et al. (2025). Genetic Susceptibility to Hidradenitis Suppurativa and Predisposition to Cardiometabolic Disease. JAMA Dermatol 161, 22–30. 10.1001/jamadermatol.2024.3779.

5. Luo, X., Ruan, Z., and Liu, L. (2024). Causal relationship between metabolic syndrome and hidradenitis suppurativa: A two-sample bidirectional Mendelian randomization study. J Dermatol 51, 1335–1349. 10.1111/1346-8138.17328.

6. Wang, H., Wu, B., Luo, M., Han, Y., Chen, J., Liu, J., Lin, L., and Xiao, X. (2024). Association of hidradenitis suppurativa (HS) with waist circumference: A bidirectional two-sample Mendelian randomization study of HS with metabolic syndrome. J Dermatol. 10.1111/1346-8138.17436.

7. Zhou, P., Jiang, X., and Wang, D. (2024). Hidradenitis suppurativa and cardiovascular diseases: A bidirectional Mendelian randomization study. Skin Res Technol 30, e13853. 10.1111/srt.13853.

8. van Straalen, K.R., Prens, E.P., Willemsen, G., Boomsma, D.I., and van der Zee, H.H. (2020). Contribution of Genetics to the Susceptibility to Hidradenitis Suppurativa in a Large, Cross-sectional Dutch Twin Cohort. JAMA Dermatol 156, 1359. 10.1001/jamadermatol.2020.3630.

9. Kjærsgaard Andersen, R., Clemmensen, S.B., Larsen, L.A., Hjelmborg, J. v. B., Ødum, N., Jemec, G.B.E., and Christensen, K. (2022). Evidence of gene–gene interaction in hidradenitis suppurativa: a nationwide registry study of Danish twins. British Journal of Dermatology 186, 78–85. 10.1111/bjd.20654.

10. Bruinsma, R.L., Fajgenbaum, K., Yang, Y., Mar Melendez-Gonzalez, M., Mohlke, K.L., Li, Y., and Sayed, C. (2021). Assessment of familial risk in patients with hidradenitis suppurativa. British Journal of Dermatology 184, 753–754. 10.1111/bjd.19664.

11. Vellaichamy, G., Dimitrion, P., Zhou, L., Ozog, D., Lim, H.W., Liao, W., Hamzavi, I.H., and Mi, Q.-S. (2021). Insights from γ-Secretase: Functional Genetics of Hidradenitis Suppurativa. Journal of Investigative Dermatology 141, 1888–1896. 10.1016/j.jid.2021.01.023.

12. Sun, Q., Broadaway, K.A., Edmiston, S.N., Fajgenbaum, K., Miller-Fleming, T., Westerkam, L.L., Melendez-Gonzalez, M., Bui, H., Blum, F.R., Levitt, B., et al. (2023). Genetic Variants Associated With Hidradenitis Suppurativa. JAMA Dermatol 159, 930. 10.1001/jamadermatol.2023.2217.

13. Kjærsgaard Andersen, R., Stefansdottir, L., Riis, P.T., Halldorsson, G., Ferkingstad, E., Oddsson, A., Walters, B., Olafsdottir, T.A., Rutsdottir, G., Zachariae, C., et al. (2024). A genome-wide association meta-analysis links hidradenitis suppurativa to common and rare sequence variants causing disruption of the Notch and Wnt/β-catenin signaling pathways. J Am Acad Dermatol. 10.1016/j.jaad.2024.11.050.

14. Gaziano, J.M., Concato, J., Brophy, M., Fiore, L., Pyarajan, S., Breeling, J., Whitbourne, S., Deen, J., Shannon, C., Humphries, D., et al. (2016). Million Veteran Program: A mega-biobank to study genetic influences on health and disease. J Clin Epidemiol 70, 214–223. 10.1016/j.jclinepi.2015.09.016.

15. Vujkovic, M., Ramdas, S., Lorenz, K.M., Guo, X., Darlay, R., Cordell, H.J., He, J., Gindin, Y., Chung, C., Myers, R.P., et al. (2022). A multiancestry genome-wide association study of unexplained chronic ALT elevation as a proxy for nonalcoholic fatty liver disease with histological and radiological validation. Nat Genet 54, 761–771. 10.1038/s41588-022-01078-z.

16. Hunter-Zinck, H., Shi, Y., Li, M., Gorman, B.R., Ji, S.-G., Sun, N., Webster, T., Liem, A., Hsieh, P., Devineni, P., et al. (2020). Genotyping Array Design and Data Quality Control in the Million Veteran Program. The American Journal of Human Genetics 106, 535–548. 10.1016/j.ajhg.2020.03.004.

17. Taliun, D., Harris, D.N., Kessler, M.D., Carlson, J., Szpiech, Z.A., Torres, R., Taliun, S.A.G., Corvelo, A., Gogarten, S.M., Kang, H.M., et al. (2021). Sequencing of 53,831 diverse genomes from the NHLBI TOPMed Program. Nature 590, 290–299. 10.1038/s41586-021-03205-y.

18. Strunk, A., Midura, M., Papagermanos, V., Alloo, A., and Garg, A. (2017). Validation of a Case-Finding Algorithm for Hidradenitis Suppurativa Using Administrative Coding from a Clinical Database. Dermatology 233, 53–57. 10.1159/000468148.

19. Mbatchou, J., Barnard, L., Backman, J., Marcketta, A., Kosmicki, J.A., Ziyatdinov, A., Benner, C., O’Dushlaine, C., Barber, M., Boutkov, B., et al. (2021). Computationally efficient whole-genome regression for quantitative and binary traits. Nat Genet 53, 1097– 1103. 10.1038/s41588-021-00870-7.

20. Willer, C.J., Li, Y., and Abecasis, G.R. (2010). METAL: fast and efficient meta-analysis of genomewide association scans. Bioinformatics 26, 2190–2191. 10.1093/bioinformatics/btq340.

21. Wang, K., Li, M., and Hakonarson, H. (2010). ANNOVAR: functional annotation of genetic variants from high-throughput sequencing data. Nucleic Acids Res 38, e164–e164. 10.1093/nar/gkq603.

22. Boyle, A.P., Hong, E.L., Hariharan, M., Cheng, Y., Schaub, M.A., Kasowski, M., Karczewski, K.J., Park, J., Hitz, B.C., Weng, S., et al. (2012). Annotation of functional variation in personal genomes using RegulomeDB. Genome Res 22, 1790–1797. 10.1101/gr.137323.112.

23. Chang, C.C., Chow, C.C., Tellier, L.C., Vattikuti, S., Purcell, S.M., and Lee, J.J. (2015). Second-generation PLINK: rising to the challenge of larger and richer datasets. Gigascience 4, 7. 10.1186/s13742-015-0047-8.

24. Machiela, M.J., and Chanock, S.J. (2015). LDlink: a web-based application for exploring population-specific haplotype structure and linking correlated alleles of possible functional variants. Bioinformatics 31, 3555–3557. 10.1093/bioinformatics/btv402.

25. Kurki, M.I., Karjalainen, J., Palta, P., Sipilä, T.P., Kristiansson, K., Donner, K.M., Reeve, M.P., Laivuori, H., Aavikko, M., Kaunisto, M.A., et al. (2023). FinnGen provides genetic insights from a well-phenotyped isolated population. Nature 613, 508–518. 10.1038/s41586-022-05473-8.

26. Ritari, J., Hyvärinen, K., Clancy, J., FinnGen, Partanen, J., and Koskela, S. (2020). Increasing accuracy of HLA imputation by a population-specific reference panel in a FinnGen biobank cohort. NAR Genom Bioinform 2, lqaa030. 10.1093/nargab/lqaa030.

27. Sernicola, A., Mazzetto, R., Tartaglia, J., Ciolfi, C., Miceli, P., and Alaibac, M. (2023). Role of Human Leukocyte Antigen Class II in Antibody-Mediated Skin Disorders. Medicina (B Aires) 59, 1950. 10.3390/medicina59111950.

28. Dedmon, L.E. (2020). The genetics of rheumatoid arthritis. Rheumatology 59, 2661–2670. 10.1093/rheumatology/keaa232.

29. Almuhanna, N., Finstad, A., and Alhusayen, R. (2021). Association between Hidradenitis Suppurativa and Inflammatory Arthritis: A Systematic Review and Meta-Analysis. Dermatology 237, 740–747. 10.1159/000514582.

30. Bizzari, S., Nair, P., Al Ali, M.T., and Hamzeh, A.R. (2017). Meta-analyses of the association of *HLA-DRB1* alleles with rheumatoid arthritis among Arabs. Int J Rheum Dis 20, 832–838. 10.1111/1756-185X.12922.

31. Said, N.M., Ezzeldin, N., Said, D., Ebaid, A.M., Atef, D.M., and Atef, R.M. (2021). HLA- DRB1, IRF5, and CD28 gene polymorphisms in Egyptian patients with rheumatoid arthritis: susceptibility and disease activity. Genes Immun 22, 93–100. 10.1038/s41435-021-00134-8.

32. Marinović, I., Čečuk-Jeličić, E., Perković, D., Marasović Krstulović, D., Aljinović, J., Šošo, D., Škorić, E., and Martinović Kaliterna, D. (2022). Association of HLA-DRB1 alleles with rheumatoid arthritis in Split-Dalmatia County in southern Croatia. Wien Klin Wochenschr 134, 463–470. 10.1007/s00508-022-02010-5.

33. van der Helm-van Mil, A.H.M., Huizinga, T.W.J., Schreuder, G.M.Th., Breedveld, F.C., de Vries, R.R.P., and Toes, R.E.M. (2005). An independent role of protective HLA class II alleles in rheumatoid arthritis severity and susceptibility. Arthritis Rheum 52, 2637–2644. 10.1002/art.21272.

34. Gregersen, P.K., Silver, J., and Winchester, R.J. (1987). The shared epitope hypothesis. an approach to understanding the molecular genetics of susceptibility to rheumatoid arthritis. Arthritis Rheum 30, 1205–1213. 10.1002/art.1780301102.

35. Kissel, T., van Schie, K.A., Hafkenscheid, L., Lundquist, A., Kokkonen, H., Wuhrer, M., Huizinga, T.W., Scherer, H.U., Toes, R., and Rantapää-Dahlqvist, S. (2019). On the presence of HLA-SE alleles and ACPA-IgG variable domain glycosylation in the phase preceding the development of rheumatoid arthritis. Ann Rheum Dis 78, 1616–1620. 10.1136/annrheumdis-2019-215698.

36. Byrd, A.S., Carmona-Rivera, C., O’Neil, L.J., Carlucci, P.M., Cisar, C., Rosenberg, A.Z., Kerns, M.L., Caffrey, J.A., Milner, S.M., Sacks, J.M., et al. (2019). Neutrophil extracellular traps, B cells, and type I interferons contribute to immune dysregulation in hidradenitis suppurativa. Sci Transl Med 11. 10.1126/scitranslmed.aav5908.

37. Mariottoni, P., Jiang, S.W., Prestwood, C.A., Jain, V., Suwanpradid, J., Whitley, M.J., Coates, M., Brown, D.A., Erdmann, D., Corcoran, D.L., et al. (2021). Single-Cell RNA Sequencing Reveals Cellular and Transcriptional Changes Associated With M1 Macrophage Polarization in Hidradenitis Suppurativa. Front Med (Lausanne) 8. 10.3389/fmed.2021.665873.

38. Zhang, M., Lin, S., Yuan, X., Lin, Z., and Huang, Z. (2019). HLA-DQB1 and HLA-DRB1 Variants Confer Susceptibility to Latent Autoimmune Diabetes in Adults: Relative Predispositional Effects among Allele Groups. Genes (Basel) 10, 710. 10.3390/genes10090710.

39. Besançon, S., Govender, D., Sidibé, A.T., Noble, J.A., Togo, A., Lane, J.A., Mack, S.J., Atkinson, M.A., Wasserfall, C.H., Kakkat, F., et al. (2022). Clinical features, biochemistry, and *HLA* - *DRB1* status in youth-onset type 1 diabetes in Mali. Pediatr Diabetes 23, 1552–1559. 10.1111/pedi.13411.

40. Zabeen, B., Govender, D., Hassan, Z., Noble, J.A., Lane, J.A., Mack, S.J., Atkinson, M.A., Azad, K., Wasserfall, C.H., and Ogle, G.D. (2019). Clinical features, biochemistry and HLA-DRB1 status in children and adolescents with diabetes in Dhaka, Bangladesh. Diabetes Res Clin Pract 158, 107894. 10.1016/j.diabres.2019.107894.

41. Fawwad, A., Govender, D., Ahmedani, M.Y., Basit, A., Lane, J.A., Mack, S.J., Atkinson, M.A., Henry Wasserfall, C., Ogle, G.D., and Noble, J.A. (2019). Clinical features, biochemistry and HLA-DRB1 status in youth-onset type 1 diabetes in Pakistan. Diabetes Res Clin Pract 149, 9–17. 10.1016/j.diabres.2019.01.023.

42. Gonzalez-Gay, M.A., Gonzalez-Juanatey, C., Lopez-Diaz, M.J., Piñeiro, A., Garcia-Porrua, C., Miranda-Filloy, J.A., Ollier, W.E.R., Martin, J., and Llorca, J. (2007). HLA–DRB1 and persistent chronic inflammation contribute to cardiovascular events and cardiovascular mortality in patients with rheumatoid arthritis. Arthritis Care Res (Hoboken) 57, 125–132. 10.1002/art.22482.

43. Farragher, T.M., Goodson, N.J., Naseem, H., Silman, A.J., Thomson, W., Symmons, D., and Barton, A. (2008). Association of the HLA–DRB1 gene with premature death, particularly from cardiovascular disease, in patients with rheumatoid arthritis and inflammatory polyarthritis. Arthritis Rheum 58, 359–369. 10.1002/art.23149.

44. Liang, W., Sun, F., Zhao, Y., Shan, L., and Lou, H. (2020). Identification of Susceptibility Modules and Genes for Cardiovascular Disease in Diabetic Patients Using WGCNA Analysis. J Diabetes Res 2020, 1–11. 10.1155/2020/4178639.

45. Paakkanen, R., Lokki, M.-L., Seppänen, M., Tierala, I., Nieminen, M.S., and Sinisalo, J. (2012). Proinflammatory HLA-DRB1*01-haplotype predisposes to ST-elevation myocardial infarction. Atherosclerosis 221, 461–466. 10.1016/j.atherosclerosis.2012.01.024.

46. Radhakrishna, U., Ratnamala, U., Jhala, D.D., Uppala, L. V, Vedangi, A., Patel, M., Vadsaria, N., Shah, S., Saiyed, N., Rawal, R.M., et al. (2023). Hidradenitis suppurativa presents a methylome dysregulation capable to explain the pro-inflammatory microenvironment: Are these DNA methylations potential therapeutic targets? J Eur Acad Dermatol Venereol 37, 2109–2123. 10.1111/jdv.19286.

47. Bao, A., Kollings, J., Ma, E., Manjunath, J., D’Amiano, A., Driscoll, M.S., and Kwatra, S.G. (2025). Association of human leukocyte antigen allelic variants with hidradenitis suppurativa across Fitzpatrick skin types: A cross-sectional analysis. J Am Acad Dermatol 92, 567–569. 10.1016/j.jaad.2024.10.031.

48. Ocejo-Vinyals, J.G., Gonzalez-Gay, M.A., Fernández-Viña, M.A., Cantos-Mansilla, J., Vilanova, I., Blanco, R., and González-López, M.A. (2020). Association of Human Leukocyte Antigens Class II Variants with Susceptibility to Hidradenitis Suppurativa in a Caucasian Spanish Population. J Clin Med 9. 10.3390/jcm9103095.

49. O’Loughlin, S., Woods, R., Kirke, P.N., Shanahan, F., Byrne, A., and Drury, M.I. (1988). Hidradenitis suppurativa. Glucose tolerance, clinical, microbiologic, and immunologic features and HLA frequencies in 27 patients. Arch Dermatol 124, 1043–1046. 10.1001/archderm.124.7.1043.

50. Montoya, M., Khetani, R., Martinez, R., Mayur, O., Yi, J.Z., and McGee, J.S. (2025). A Genome-Wide Survey of DNA Methylation Status in Whole Blood of Patients With Hidradenitis Suppurativa Suggests Systemic Immune Dysregulation and Systemic Disease Burden. Exp Dermatol 34, e70065. 10.1111/exd.70065.

51. Vidal, V.P.I., Chaboissier, M.-C., Lützkendorf, S., Cotsarelis, G., Mill, P., Hui, C.-C., Ortonne, N., Ortonne, J.-P., and Schedl, A. (2005). Sox9 Is Essential for Outer Root Sheath Differentiation and the Formation of the Hair Stem Cell Compartment. Current Biology 15, 1340–1351. 10.1016/j.cub.2005.06.064.

52. Kadaja, M., Keyes, B.E., Lin, M., Pasolli, H.A., Genander, M., Polak, L., Stokes, N., Zheng, D., and Fuchs, E. (2014). SOX9: a stem cell transcriptional regulator of secreted niche signaling factors. Genes Dev 28, 328–341. 10.1101/gad.233247.113.

53. Nowak, J.A., Polak, L., Pasolli, H.A., and Fuchs, E. (2008). Hair Follicle Stem Cells Are Specified and Function in Early Skin Morphogenesis. Cell Stem Cell 3, 33–43. 10.1016/j.stem.2008.05.009.

54. Luo, H., Liu, W., Zhou, Y., Zhang, Y., Wu, J., Wang, R., and Shao, L. (2022). Stage-specific requirement for METTL3-dependent m6A modification during dental pulp stem cell differentiation. J Transl Med 20, 605. 10.1186/s12967-022-03814-9.

55. Orvain, C., Lin, Y.-L., Jean-Louis, F., Hocini, H., Hersant, B., Bennasser, Y., Ortonne, N., Hotz, C., Wolkenstein, P., Boniotto, M., et al. (2020). Hair follicle stem cell replication stress drives IFI16/STING-dependent inflammation in hidradenitis suppurativa. Journal of Clinical Investigation 130, 3777–3790. 10.1172/JCI131180.

56. Place, E., Manning, E., Kim, D.W., Kinjo, A., Nakamura, G., and Ohyama, K. (2022). SHH and Notch regulate SOX9+ progenitors to govern arcuate POMC neurogenesis. Front Neurosci 16. 10.3389/fnins.2022.855288.

57. Sun, J., Chen, Q., and Ma, J. (2022). Notch–Sox9 Axis Mediates Hepatocyte Dedifferentiation in KrasG12V-Induced Zebrafish Hepatocellular Carcinoma. Int J Mol Sci 23, 4705. 10.3390/ijms23094705.

58. Haller, R., Schwanbeck, R., Martini, S., Bernoth, K., Kramer, J., Just, U., and Rohwedel, J. (2012). Notch1 signaling regulates chondrogenic lineage determination through Sox9 activation. Cell Death Differ 19, 461–469. 10.1038/cdd.2011.114.

59. Fan, X.-D., Zheng, H.-B., Fan, X.-S., and Lu, S. (2018). Increase of SOX9 promotes hepatic ischemia/reperfusion (IR) injury by activating TGF-β1. Biochem Biophys Res Commun 503, 215–221. 10.1016/j.bbrc.2018.06.005.

60. Scharf, G.M., Kilian, K., Cordero, J., Wang, Y., Grund, A., Hofmann, M., Froese, N., Wang, X., Kispert, A., Kist, R., et al. (2019). Inactivation of Sox9 in fibroblasts reduces cardiac fibrosis and inflammation. JCI Insight 4. 10.1172/jci.insight.126721.

61. Hoshino, Y., Kurabayashi, M., Kanda, T., Hasegawa, A., Sakamoto, H., Okamoto, E., Kowase, K., Watanabe, N., Manabe, I., Suzuki, T., et al. (2000). Regulated Expression of the BTEB2 Transcription Factor in Vascular Smooth Muscle Cells. Circulation 102, 2528– 2534. 10.1161/01.CIR.102.20.2528.

62. Min, C., Kirsch, K.H., Zhao, Y., Jeay, S., Palamakumbura, A.H., Trackman, P.C., and Sonenshein, G.E. (2007). The Tumor Suppressor Activity of the Lysyl Oxidase Propeptide Reverses the Invasive Phenotype of Her-2/neu–Driven Breast Cancer. Cancer Res 67, 1105–1112. 10.1158/0008-5472.CAN-06-3867.

63. Lyu, Y., Guan, Y., Deliu, L., Humphrey, E., Frontera, J.K., Yang, Y.J., Zamler, D., Kim, K.H., Mohanty, V., Jin, K., et al. (2022). KLF5 governs sphingolipid metabolism and barrier function of the skin. Genes Dev 36, 822–842. 10.1101/gad.349662.122.

64. Dany, M., and Elston, D. (2017). Gene expression of sphingolipid metabolism pathways is altered in hidradenitis suppurativa. J Am Acad Dermatol 77, 268–273.e6. 10.1016/j.jaad.2017.03.016.

65. Rosi, E., Fastame, M.T., Silvi, G., Guerra, P., Nunziati, G., Di Cesare, A., Scandagli, I., Ricceri, F., and Prignano, F. (2022). Hidradenitis Suppurativa: The Influence of Gender, the Importance of Trigger Factors and the Implications for Patient Habits. Biomedicines 10, 2973. 10.3390/biomedicines10112973.

