## Supplemental Materials for "A genome-wide association study of hidradenitis suppurativa from the VA’s Million Veteran Program"

**Supplemental Table 1**. Cardiovascular comorbidities by ancestry

|  | African American  N = 112,279 | | | European American  N = 437,982 | | | Hispanic American  N = 47,588 | | |
| --- | --- | --- | --- | --- | --- | --- | --- | --- | --- |
|  | HS  N = 1,931 | Non- HS  N = 110,348 | p-value | HS  N = 2,613 | Non- HS  N = 435,369 | p-value | HS  N = 415 | Non-HS  N = 47,173 | p-value |
| Hypertension | 82.3% | 82.1% | 0.82 | 81.4% | 77.2% | <0.001 | 69.4% | 66.3% | 0.21 |
| Hyperlipidemia | 75.1% | 75.3% | 0.87 | 83.7% | 80.9% | <0.001 | 76.1% | 75.8% | 0.007 |
| Type 1 diabetes | 15.6% | 10.4% | <0.001 | 14.4% | 7.4% | <0.001 | 15.4% | 8.5% | <0.001 |
| Type 2 diabetes | 53.7% | 48.4% | <0.001 | 52.0% | 39.2% | <0.001 | 54.5% | 43.2% | <0.001 |
| Myocardial infarction | 12.5% | 11.0% | 0.038 | 17.8% | 13.6% | <0.001 | 12.0% | 9.2% | 0.063 |
| Cerebrovascular accident | 23.9% | 21.1% | 0.004 | 28.4% | 23.5% | <0.001 | 20.5% | 16.8% | 0.051 |

**Supplemental Table 2**. Confirmation of MVP results using FinnGen, UK Biobank and HS ProCARE

|  | Chr. | pos_hg38 | A1 | A2 | EA | beta | se | p-value | EAF | Note |
| --- | --- | --- | --- | --- | --- | --- | --- | --- | --- | --- |
| EA | 1 | 64647885 | T | C |  |  |  |  |  | Absent in all the three studies |
| EA | 1 | 70340139 | G | A | A | -0.012 | 0.096 | 0.9 | 0.0174 | AF in UKB 4%, in FinnGen 0.7%, absent int our data. Different directions in UKB and FinnGen. UKB p-value 0.16 with A increasing disease risk; FinnGen p-value 0.26 |
| HA | 6 | 159231273 | GCTGAGGCAGGAGA | G |  |  |  |  |  | absent in all the three studies |
| HA | 9 | 108737932 | T | C | T | 0.07 | 0.1 | 0.4894 | 0.9846 | AF in UKB 96.4%, p-value 0.74; AF in FinnGen 99.4%, p-value 0.53; same direction. |
| META | 6 | 32603181 | T | G | T | 0.074 | 0.034 | 0.03 | 0.1625 | AF in UKB 18.6%, p-value 0.63; AF in FinnGen 0.152, p-value 0.025; same direction. |
| META | 17 | 71524020 | T | C | T | 0.149 | 0.028 | 7.50E-08 | 0.1309 | This variant in high LD (R2 = 0.966 in TOP-LD EUR) with our lead variant chr17:71515958:G:A |

Abbreviations: AA, African ancestry; EA, European ancestry; FREQ, frequency; HA, Hispanic ancestry; META, meta-analysis, STD ERR, standard error

**Supplemental Figure 1**. HS GWAS of AA subjects (n=1,931)

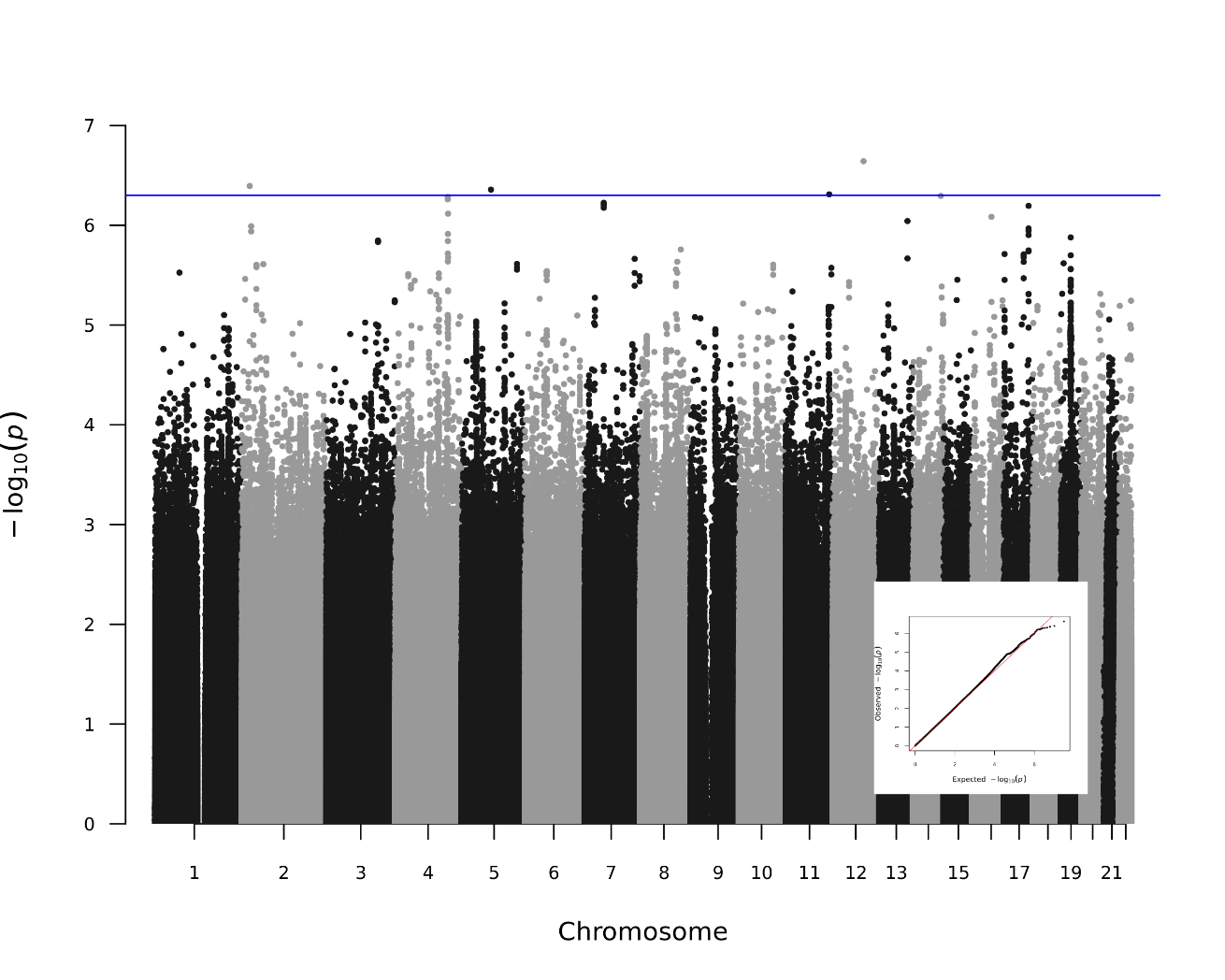

**Supplemental Figure 2. HS GWAS of EA subjects (n=2,613)**

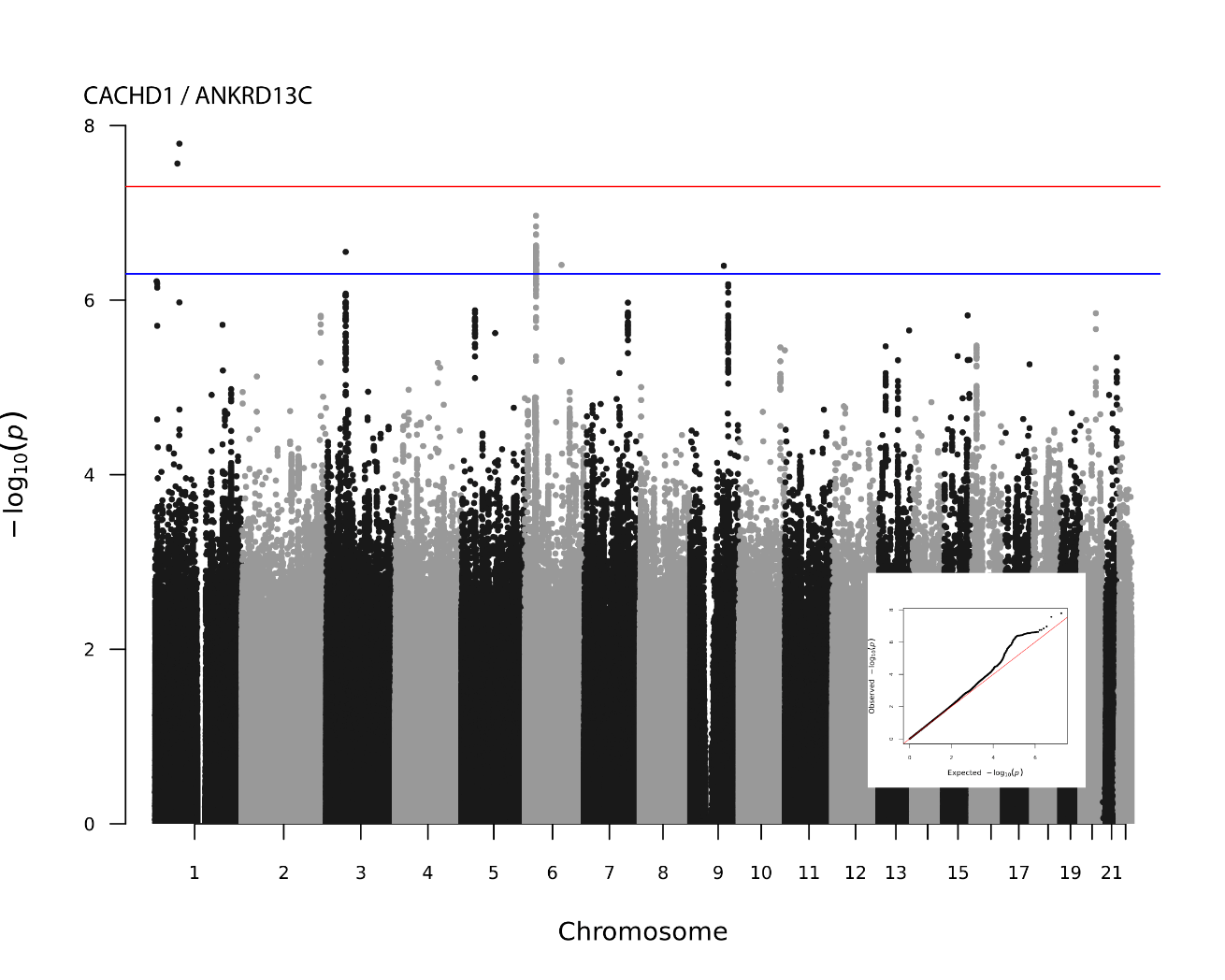

**Supplemental Figure 3.** HS GWAS of HA subjects (n=415)

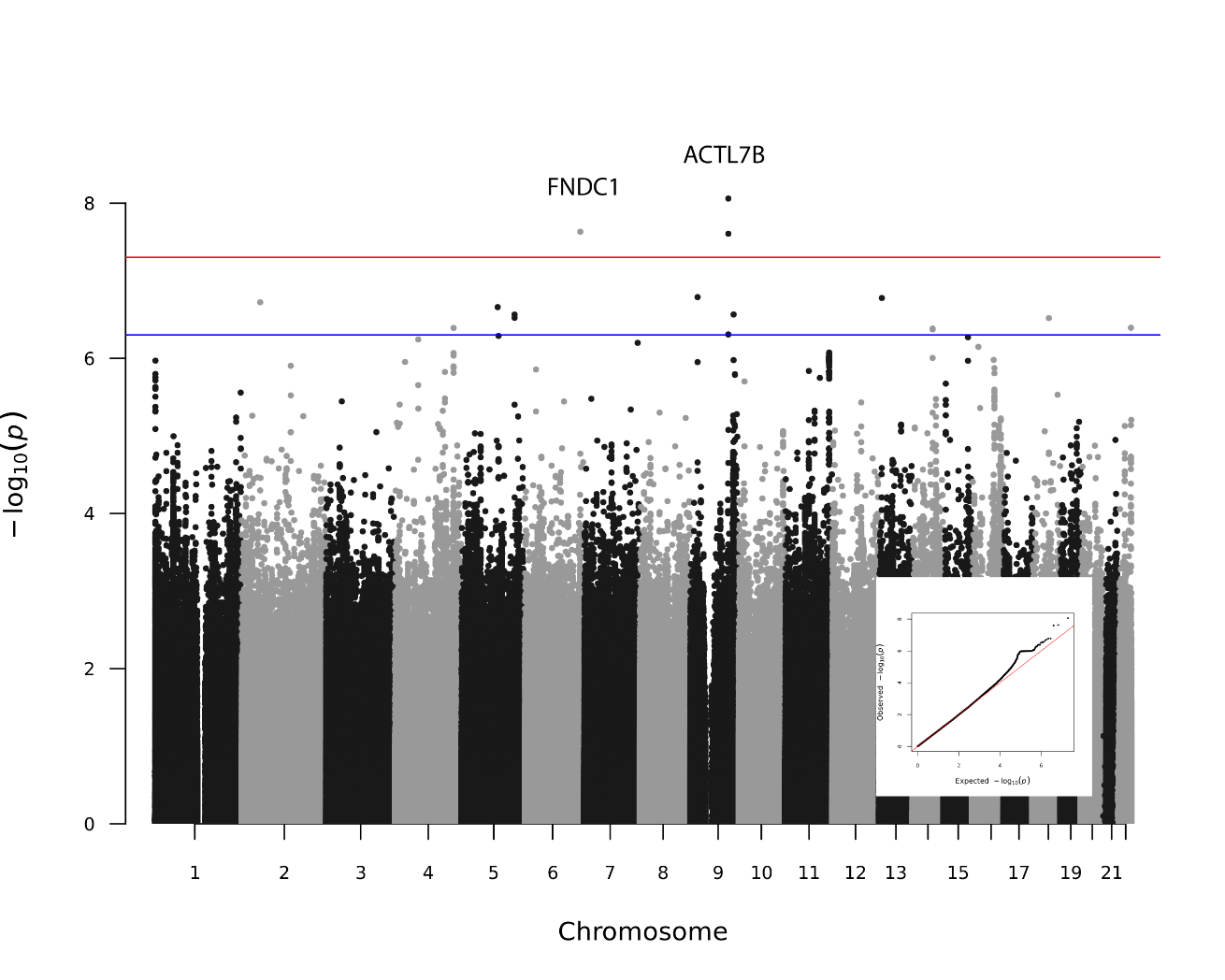
